# Polycystic ovary syndrome susceptibility loci inform disease etiological heterogeneity

**DOI:** 10.1101/2021.03.04.21252927

**Authors:** Yanfei Zhang, Vani C. Movva, Marc S. Williams, Ming Ta Michael Lee

## Abstract

**Purpose:** Polycystic ovary syndrome (PCOS) is a complex disorder with heterogenous phenotypes and unclear etiology. A recent phenotypic clustering study identified metabolic and reproductive subtypes of PCOS. We attempted to deconstruct the PCOS heterogeneity from a genetic perspective.

**Methods:** We applied *k-means* clustering to categorize the genome-wide significant PCOS variants into clusters based on their associations with selected quantitative traits that likely reflect PCOS etiological pathways. We evaluated the association of each cluster with PCOS related traits and disease outcomes. We then applied Mendelian randomization to estimate the causal effect of the traits on PCOS and PCOS on disease outcomes.

**Results:** Clustering analysis suggested three categories of variants: adiposity, insulin resistant, and reproductive. Significant associations were observed for variants in the adiposity cluster with body mass index (BMI), waist circumference and breast cancer, and variants in insulin resistant cluster with fasting insulin and glucose values, and homeostatic model assessment of insulin resistance (HOMA-IR). Sex hormone binding globulin (SHBG) has strong association with all three clusters. Mendelian randomization supported the causal role of BMI and SHBG on PCOS. No causal associations were observed for PCOS on disease outcomes.

**Main Conclusions:** Our study provides genetic evidence for the heterogeneity in PCOS etiologies, corresponding to the reported phenotypic subtypes. Such studies will improve the current PCOS diagnosis criteria that do not distinguish the heterogeneity. Classification of women with PCOS to inform appropriate treatment will be more accurate in the future with improvements in clustering analysis for PCOS.

## Introduction

Polycystic ovary syndrome (PCOS) is a complex disorder affecting approximately 15% of women of reproductive age [1]. PCOS includes highly heterogeneous phenotypic manifestations characterized by a variety of reproductive and metabolic abnormalities, including ovulatory dysfunction, hyperandrogenism, hirsutism, obesity, and insulin resistance [1]. The commonly used National Institutes of Health (NIH) [2] and Rotterdam [3, 4] diagnostic criteria for PCOS are designed to account for the diverse phenotypic presentations but do not provide mechanistic insights [5]. The etiology or etiologies of PCOS are still unclear.

To obtain insight on etiology and deconstruct the heterogeneity of PCOS, a recent study performed clustering analysis using body mass index (BMI) and seven biochemical biomarkers in a PCOS cohort and identified two distinct phenotypic clusters: a “reproductive” subtype characterized by high luteinizing hormone (LH) and sex hormone binding globulin (SHBG) levels with low BMI and insulin levels, and a “metabolic” subtype characterized by high BMI, glucose, and insulin levels with low SHBG and LH levels [5]. It is important to note that biochemical markers change with many factors such as aging of a person, usage of insulin and contraceptive pills. It is also unclear whether these biomarkers are causal or consequential to the disease. Unlike biomarkers, germline DNA remains constant regardless of external factors and age. Genetic variants are often used as an instrument to explore causality.

PCOS is highly heritable with an estimated heritability of 38-71% as noted in the twin study [6]. Recent large-scale genome-wide studies (GWAS) brought significant progress in identification of PCOS susceptibility loci [7-12]. Thirty-seven variants with genome-wide significance have been identified so far by GWAS in European and East Asian populations, offering insights into causal biological pathways for PCOS. With the public access to GWAS dataset of many traits and disease outcomes, it is now possible to elucidate disease mechanisms using variant clustering techniques assuming that genetic variants that act along a shared pathway will have similar directional effect on a trait [13]. Such a strategy was previously applied to deconstruct the mechanistic heterogeneity of type 2 diabetes mellitus (T2DM) [13-15].

In this study, we hypothesize that the heterogeneity of PCOS manifestations reflects different mechanistic pathways and can be identified using a genetic approach. We performed clustering analysis on the association of PCOS susceptibility loci with various traits that are related to PCOS. We then used genetic risk scoring to evaluate the effect of each cluster. We also performed Mendelian randomization to estimate the causal effect of the traits on PCOS and PCOS on disease outcomes.

## Methods

### Selection of PCOS associated genetic variants, traits and disease outcomes

We compiled a list of 37 genome-wide significant variants associated with PCOS (p<5e-8) from previously published GWAS (Table S1). Only variants that are not in linkage disequilibrium (LD R^2^<0.5) were included. Four variants were excluded later as they were not included in the summary statistics of most of the traits or disease outcomes, and no proxy single nucleotide variants (SNVs) could be identified for them. A total of 26 variants were included in the final analysis. The rs10993397 (*C9orf3*) and rs8043701 (*TOX3*) were replaced by their proxy SNVs rs7865239 and rs11075468 (Table S1).

We selected four groups of traits that are likely to reflect PCOS etiologies: 1) adiposity traits: female body mass index (BMI, general adiposity), female waist circumstances (WC) and female waist hip ratio (WHR, central adiposity) [16, 17]; 2) hormonal traits: sex hormone binding globulin (SHBG), luteinizing hormone (LH) [18]; 3) insulin resistant traits: fasting insulin (FI), fasting glucose (FG), homeostatic model assessment of insulin resistance (HOMA-IR) [19, 20]; 4) lipids: high-density lipoprotein (HDL), low-density lipoprotein (LDL), total cholesterol (TC) and triglycerides (TG)[21]. Disease outcomes include T2DM [22], coronary artery disease (CAD) [23], and breast cancer[24]. Traits and outcomes like follicle stimulating hormone (FSH), testosterone, dehydroepiandrosterone sulfate (DEHAS), other female reproductive organ cancers such as endometrial and ovarian cancer, although planned, could not be included as the GWAS datasets were unavailable or included very few PCOS susceptibility loci and proxy SNVs. The TwoSampleMR package was development to ease Mendelian randomization analysis [25]. It connects to the IEU open GWAS database and make it convenient to extract and harmonize data. We employed TwoSampleMR package to retrieve, read in and harmonize the association summary statistics of PCOS variants with these traits and outcomes (datasets listed in supplementary table 2).

### Clustering

First, we calculated the z-score (z-score = β/se) from the summary statistics of the 26 PCOS variants from the GWAS of the four groups of quantitative traits (supplementary table 3). All effects were aligned to the PCOS risk-increasing alleles. We then applied *k-means* clustering method on the association z-scores where variants are clustered together based on the similar associations with the traits. This method is widely applied on quantitative data and was successfully used by our team to identify subgroups of patients with different responses to phenylephrine [26]. As *k-means* clustering requires the number of clusters in advance, we used the NbClust package to decide the best number of clusters [27]. Supplementary figure 1 showed that 8 indices suggest a three-cluster solution. Analyses were performed in R (3.6.3).

**Figure 1.**
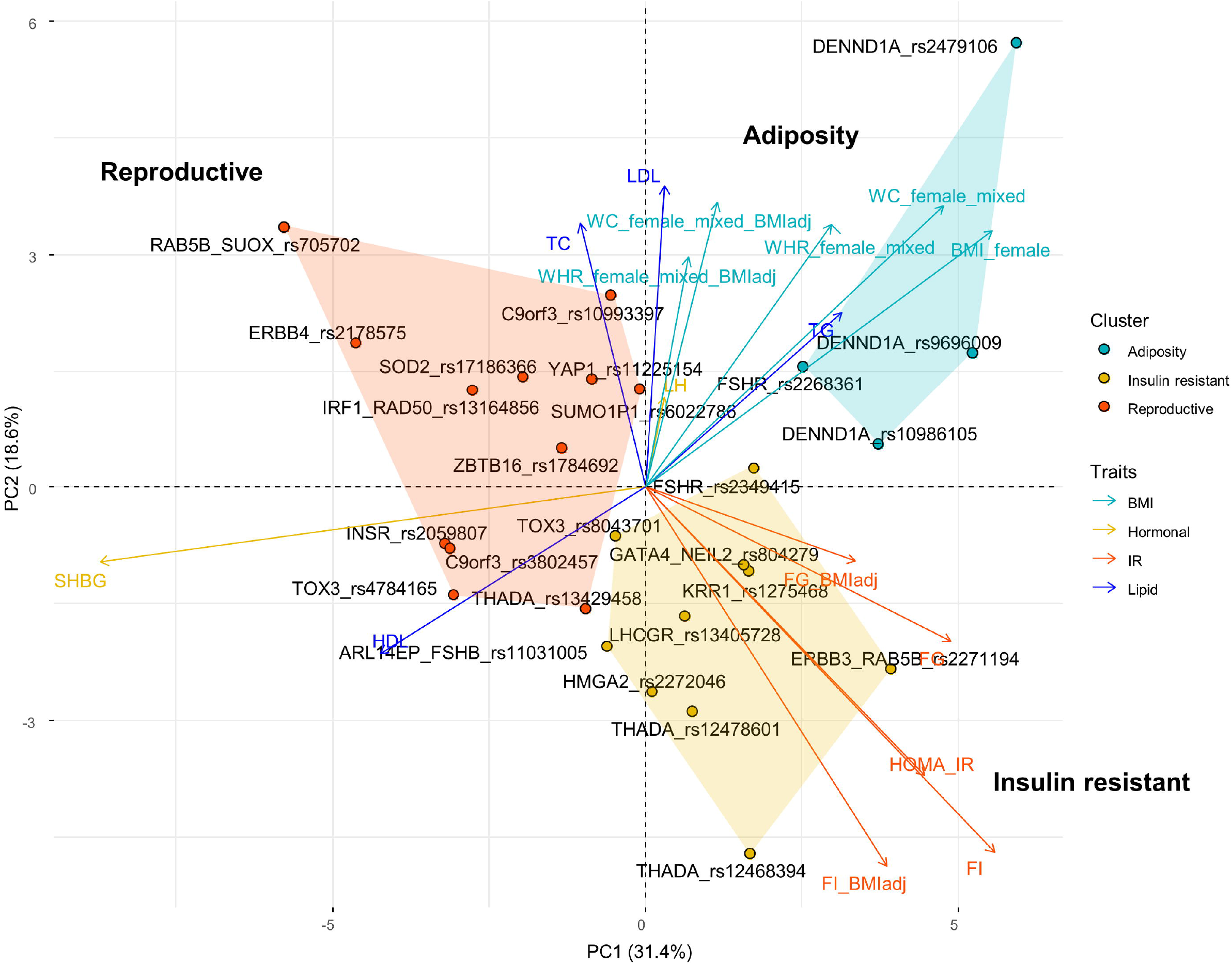
PCA plot of the variant-trait associations for PCOS variants. PCOS variants are plotted on the first 2 principal components (PCs) of the association Z-score and colored by the assigned clusters. The relative magnitude and direction of trait correlation with the PCs are shown with arrows. BMI, body mass index; WC: waist circumference; WHR: waist hip ratio; PC, principal component; PCA, principal component analysis; FG: fasting glucose; FI: fasting insulin; SHBG, sex hormone binding globulin; LH, luteinizing hormone; HDL: high-density lipoprotein; LDL: low-density lipoprotein; TC: total cholesterol; TG: triglycerides.

### Trait and disease associations with each cluster

The association of the genetic risk scores of each cluster with each trait and disease outcome was performed by an inverse-variance fixed effect meta-analysis of the summary statistics of the variant-trait and -disease from GWAS described previously [13-15]. Association with five disease outcomes, which were not used in clustering analysis were also examined. P-value<0.0024 was considered significant with Bonferroni correction for 16 traits and 5 disease outcomes (0.05/21).

### Mendelian randomization analysis

Based on the association results of genetic risk score with traits and disease outcomes, we performed Mendelian randomization analysis to evaluate the causal role of SHBG, BMI, WC, WHR, and insulin resistance on PCOS. Instrumental variables for SHBG, BMI, WC, WHR were extracted from curated dataset by the IEU open GWAS project (supplementary table 2). For insulin resistance, we used the 53 significant variants associated with an integrated insulin resistant phenotype composed of FI, TG, and HDL [28]. We adopted the β and SE from study of Wang *et al*., who meta-analyzed the absolute value of the standardized β coefficient for each of the 53 SNV associations with the individual components of the composite IR phenotype using a fixed-effect inverse-variance weighted (IVW) method (supplementary table 4)[29]. For the PCOS outcome data, we meta-analyzed the results from studies by Day *et al*., (without samples from 23andme) [11] and Zhang *et al*., [12] using METAL [30]. When PCOS is an exposure, we use the 14 variants reported in so far, the largest meta-analysis as instrumental variables [11]. TwoSampleMR R package were used to perform MR using inverse variance weighted method and sensitivity analysis using MR-Egger and Weighted median methods [25].

## Results

### Clustering suggests mechanistic heterogeneity for PCOS etiology

Clustering of variant-trait associations using 26 PCOS variants and 16 traits identified 3 clusters of variants. Three clusters were mainly distinguished by the BMI related traits, insulin resistant traits, and SHBG, as visualized in the PCA plot (Figure 1). Thus, we named them “adiposity”, “insulin resistant” and “reproductive” clusters. Table 1 lists the associations of genetic risk score of each cluster and the traits. Four variants were included in adiposity cluster, including three variants in *DENND1A* and a variant in *FSHR*. BMI (β = 0.015, p= 2.59e-7) and WC (β = 0.017, p =1.67e-7) are the most significantly associated traits. Even with adjustment of BMI, WC remains significant (β=0.01, p=0.001). WHR is only significant without BMI adjustment (β = 0.011, p = 0.0008). Additionally, the adiposity cluster is negatively associated with SHBG (β = - 0.198, p=7.34e-6). The insulin resistant cluster has 10 variants associated with *THADA, LHCGR, FSHB, FSHR, ERBB3, TOX3, GATA4*, and *KRR1*. The most significant trait is fasting insulin (β=0.005, p=1.93e-5), and the same was true with BMI adjustment as well (β=0.004, p=2.12e-5). HOMA-IR is also significantly associated (β = 0.005, p=0.0013). Fasting glucose is only significant without BMI adjustment (β = 0.004, p = 0.0004). Another significant trait is total cholesterol (β = −0.006, p= 0.0009). The reproductive cluster includes variants in genes of *ERBB4, RAB5B, IRF1, SOD2, YAP1, SUMO1B, ZBTB, C9orf3, INSR, THADA*, and *TOX3*. SHBG is the only significant trait associated with this cluster (β= 0.0052, p=1.70e-6).

**Table 1.** The association of the genetic risk score of each cluster with traits and disease outcomes

|  | Adiposity |  | Insulin resistant |  | Reproductive |  |
| --- | --- | --- | --- | --- | --- | --- |
| | $\beta$ | p-value | $\beta$ | p-value | $\beta$ | p-value |
| SHBG | -0.198 | 7.377E-06 | -0.069 | 0.0052 | 0.111 | 1.70E-06 |
| LH | 0.025 | 0.1462 | 0.002 | 0.8053 | -0.001 | 0.8898 |
| FI | 0.004 | 0.0855 | 0.005 | 1.93E-05 | -0.003 | 0.0101 |
| FI-BMI adjusted | 0.002 | 0.3702 | 0.004 | 2.12E-05 | -0.001 | 0.1558 |
| FG | 0.004 | 0.0519 | 0.004 | 0.0004 | -0.002 | 0.0653 |
| FG-BMI adjusted | 0.003 | 0.1947 | 0.003 | 0.0138 | -0.001 | 0.3067 |
| HOMA-IR | 0.003 | 0.2389 | 0.005 | 0.0013 | -0.003 | 0.0633 |
| HDL | -0.008 | 0.0035 | 0 | 0.9734 | 0.003 | 0.0411 |
| LDL | 0.003 | 0.335 | -0.004 | 0.0152 | -0.002 | 0.2336 |
| TG | 0.001 | 0.6019 | -0.003 | 0.0499 | -0.004 | 0.0207 |
| TC | -0.001 | 0.7885 | -0.006 | 0.0009 | -0.001 | 0.5062 |
| BMI | 0.015 | 2.59E-07 | 0 | 0.9586 | -0.001 | 0.4855 |
| WC | 0.017 | 1.67E-07 | 0 | 0.892 | 0.001 | 0.7129 |
| WC-BMI adjusted | 0.01 | 0.001 | 0 | 0.8755 | 0.004 | 0.0264 |
| WHR | 0.011 | 0.0008 | -0.002 | 0.3264 | 0 | 0.8949 |
| WHR-BMI adjusted | 0.006 | 0.0807 | -0.002 | 0.251 | 0.001 | 0.5162 |
| CAD | -0.007 | 0.1482 | 0 | 0.8988 | 0 | 0.8444 |
| T2DM | 0.009 | 0.0726 | -0.004 | 0.2279 | -0.005 | 0.0964 |
| Breast Cancer | 0.014 | 0.0008 | 0.007 | 0.0106 | 0.006 | 0.0192 |
| ER positive | 0.012 | 0.0201 | 0.008 | 0.0146 | 0.005 | 0.0864 |
| ER negative | 0.016 | 0.0423 | 0.002 | 0.6789 | 0.003 | 0.4304 |

WC: waist circumference; WHR: waist-hip ratio; BC: breast cancer; ER: estrogen receptor; T2DM: type 2 diabetes mellitus; CAD: coronary artery disease; SHBG: sex hormone binding globulin; BMI: body mass index; LH: luteinizing hormone; HOMA-IR: homeostatic model assessment of insulin resistance; FI: fasting insulin; FG: fasting glucose; HDL: high-density lipoprotein; LDL: low-density lipoprotein; TG: triglyceride; TC: total cholesterol.

We also investigated the association of clusters with disease outcomes (Table 1). None of the three clusters are associated with CAD or T2DM. The adiposity cluster is significantly associated with breast cancer (β = 0.014, p = 0.0008). The insulin resistant and reproductive clusters are associated with breast cancer at nominal significance (p<0.05).

### Mendelian randomization predicts a causal role of SHBG and BMI on PCOS

The clustering analysis suggest that BMI, insulin resistance, and SHBG are involved in PCOS etiology. Thus, we employed MR to estimate the causal roles of these factors on PCOS. The inverse variance weighted method suggests causal effect of SHBG, BMI, WC and insulin resistance (Table 2). In the sensitivity analysis using weighted median and MR-Egger methods, only SHBG and BMI remained associated with causality (p<0.05). We also evaluated the causal effect of PCOS on disease outcomes. We did not observe significant evidence to support a causal role for PCOS on T2DM or CAD. There may be a causal effect of PCOS on breast cancer (β = 0.0646, p= 0.00195) especially the estrogen receptor (ER) positive type (β = 0.0862, p=7.9e-3), according to the IVW method. However, MR-Egger result is not significant, suggesting a pleiotropic effect of the instrumental variants.

**Table 2:**
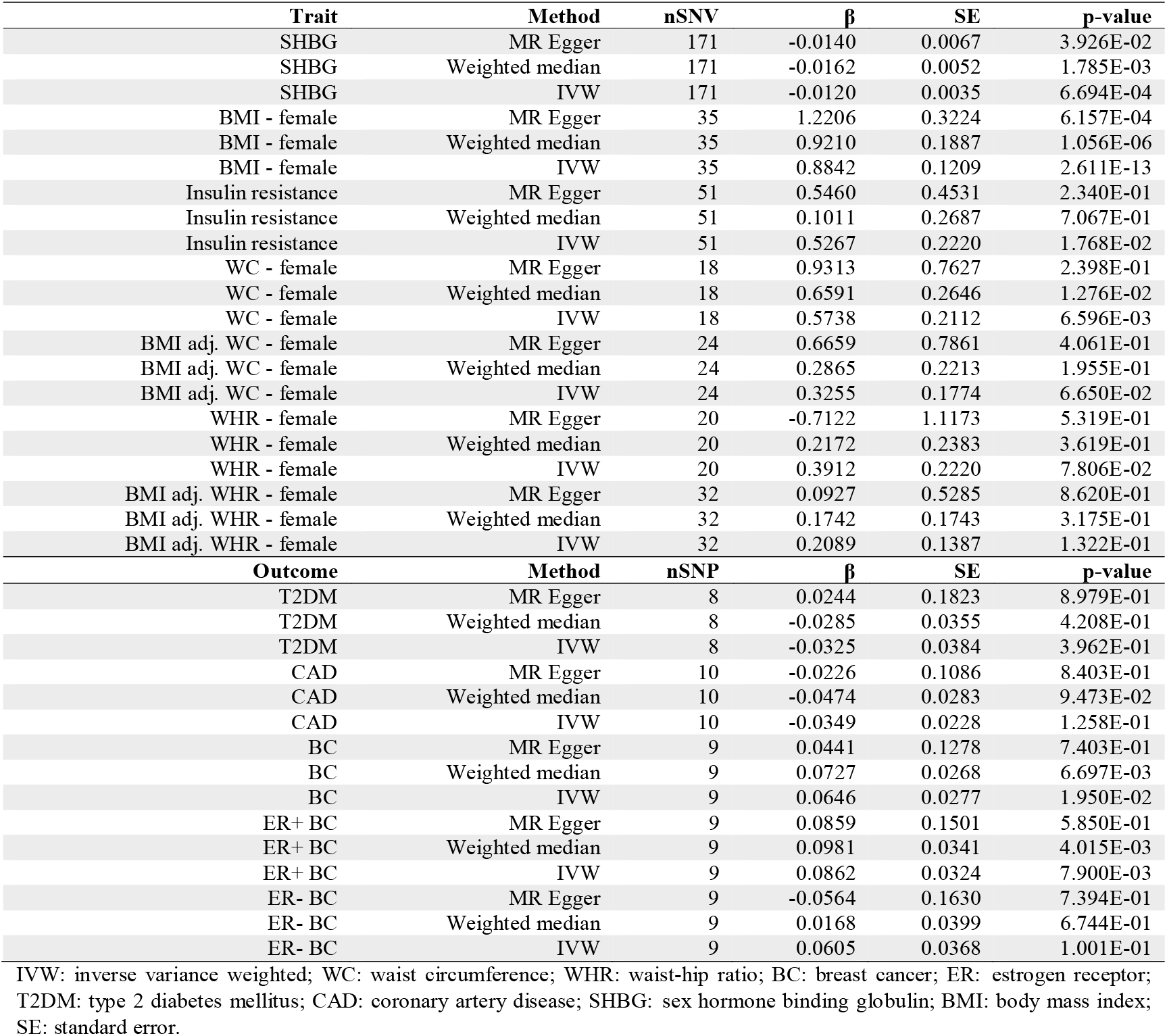
Result of Mendelian randomization of traits on PCOS and PCOS on disease outcomes

## Discussion

Common complex diseases consist of combinations of symptoms and phenotypes that may be result from different mechanistic pathways. Several studies have identified subtypes of T2DM using biomarkers or phenotypes [31, 32]. These subtypes indicate different mechanistic pathways of T2DM which were later supported by deconstruction of T2DM susceptibility loci [13, 15]. Our study performed a clustering analysis on PCOS susceptibility variants and identified three clusters of variants that associated with adiposity, insulin resistant, and hormonal traits, providing genetic evidence for the recently reported metabolic and reproductive subtypes of PCOS in a phenotypic clustering using BMI and 7 serum biochemical markers [5].

As in the phenotypic clustering study where BMI, fasting insulin, and SHBG are the key features separating the metabolic and reproductive subtypes [5], the clusters of variants in our analysis were mainly differentiated by associations with SHBG, BMI, and fasting insulin. However, BMI and fasting insulin clustered together in the metabolic subtype and cannot be separated by phenotypic clustering. In our analysis, variants in adiposity and insulin resistant clusters are clearly separated by associations with BMI and fasting insulin. In the phenotypic clustering, the metabolic subtype showed relatively higher BMI, fasting insulin, and lower SHBG while the reproductive subtype showed higher SHBG but lower BMI, and fasting insulin. In our analysis, the adiposity and insulin resistant clusters showed significant positive associations with BMI and fasting insulin but negative association with SHBG, while the reproductive cluster showed significant positive association with SHBG and negative association with BMI (not significant, p=0.4855) and fasting insulin (p=0.01). Our study also investigated WC and WHR to represent the fat distribution pattern. WC is significantly associated with adiposity cluster even after BMI adjustment. However, the association with WHR became nonsignificant with BMI adjustment. These results suggest that general adiposity, but not central adiposity, is the main associated feature. In the further analysis using MR, we confirmed the causal association of SHBG and BMI on PCOS by IVW and MR Egger method, which are consistent to previous studies[9, 33, 34]. In terms of central adiposity, the effect of WC is mainly mediated through BMI and WHR is not causally related to PCOS. We used the 53 variants associated with an integrated insulin resistant phenotype that included 3 components – high levels of fasting insulin and TG, low levels of HDL [28]. IVW method showed a causal relationship of insulin resistance and PCOS, while MR egger test suggested horizontal pleiotropy of the instrumental variables. It is possible that these variants also impact BMI.

We also explored the association of variant clusters with the disease outcome. Epidemiology studies showed increased risk of diabetes for PCOS patients [35, 36]. However, epidemiology studies often suffer from unknown confounding factors making it difficult to infer causality. In our analysis, we do not see significant associations of clusters of variants and T2DM. MR also confirmed there is no causal effect of PCOS on T2DM. Additionally, conflicting results were observed between PCOS and breast cancer [37]. In general, studies to date have not observed an association between PCOS and breast cancer risk [38]. In our study, we saw marginal associations of breast cancer with three clusters of variants. The possible causal effect of PCOS on breast cancer by MR IVW method is possibly mediated by other factors as suggested by MR-Egger.

Our study has limitations. We used k-means clustering, which is a “hard-clustering” method where one variant is assigned to only one cluster. However, this does not reflect the reality most of the times as a single gene can be involved in multiple pathways (pleiotropy). In our analysis, we observed that different variants from the same genes are classified into different clusters, such as variants in *THADA, FSHR*, and *TOX3*. Our study is also limited by the GWAS datasets available for PCOS-related traits, especially gonadotropin. For example, we did not include FSH as the GWAS dataset in our analysis has very few variants. Also, we do not see an association with LH in the variant clustering as observed in the phenotypic clustering. FSH and LH vary significantly between individuals and changes with menstrual cycle [39]. The GWAS of LH with small sample size may lack statistical power to capture the variabilities in the general population. This may explain why variants in *FSHR, FSHB*, and *LHCGR* do not cluster together. GWAS of PCOS-related disease outcomes such as hirsutism, endometrial and ovarian cancer are not available or have a limited number of variants, thus the associations of genetic risk score of each cluster with these outcomes are lacking. With increasing access to more large-scale GWAS of more granular features, future studies can apply a more sophisticated “soft-clustering” method to deconstruct the PCOS genetic architecture.

In conclusion, clustering of variants associated with PCOS has identified variant clusters of significant associations with adiposity, insulin resistance and SHBG, likely to represent the etiologies for PCOS. Subsequent MR analysis predicts a causal role for BMI and SHBG and a suggestive causal effect of insulin resistance on PCOS. Our study is the first to use a genetic approach to deconstruct PCOS etiological heterogeneity. It provides genetic evidence on the heterogeneity in PCOS etiologies, corresponding to the results of the phenotypic clustering. Such studies will accelerate the deep phenotyping of PCOS and will complement the fact that current PCOS diagnostic criteria cannot distinguish the subtypes. With progress of clustering analysis on PCOS phenotype and genetics, it is possible in future to classify women with PCOS and apply precise treatment.

## Data Availability

The data used in this study are from public dataset and the references were provided in the method and supplementary notes.

## Acknowledgement

The authors express their gratitude to the IEU open GWAS project (https://gwas.mrcieu.ac.uk/) and the consortium and teams that shared the GWAS summary statistics.

## Funding

The authors did not receive funding for this project.

## Authors’ contribution

YZ: Study conceptualization, data analysis, interpretation and drafting. VM and MTML: data interpretation and critical review. MSW: critical review.

## Notes

### Competing Interest Statement

The authors have declared no competing interest.

### Author Declarations

This study is exempted from an IRB review because only summary data were used.

