## Supplementary tables1,2,4 and figure s1 for "Polycystic ovary syndrome susceptibility loci inform disease etiological heterogeneity"

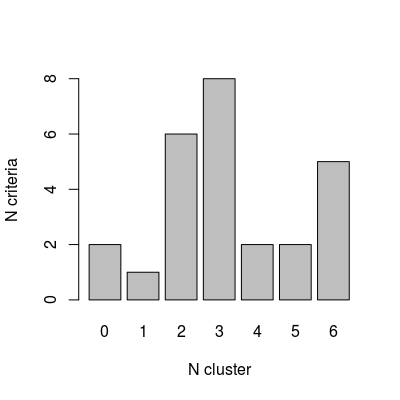

Supplementary figure 1. Using NbClust to identify the best number of clusters for k-means clustering. X-axis represents the number of clusters; y-axis represents of the number of criteria to support the number of clusters by NbClust package.

Supplementary table 1. PCOS significant variants by GWAS

| **SNV** | **CHR:BP (GRCh37)** | **Risk**  **Allele** | **Other Allele** | **Cluster** | **Gene** | **Population** | **Comment** | **PMID** |
| --- | --- | --- | --- | --- | --- | --- | --- | --- |
| rs10986105 | 9:126549955 | C | A | 1 | *DENND1A* | Han Chinese | Keep | 21151128 |
| rs10993397 | 9:97679771 | T | C | 3 | *C9orf3* | European | Keep. Using proxy SNP rs7865239 | 26284813 |
| rs11031005 | 11:30226356 | C | T | 2 | *ARL14EP/FSHB* | European | Keep | 30566500 |
| rs11225154 | 11:102043240 | A | G | 3 | *YAP1* | European | Keep | 26416764; 30566500 |
| rs12468394 | 2:43561161 | C | A | 2 | *THADA* | Han Chinese | Keep | 21151128 |
| rs12478601 | 2:43721508 | C | T | 2 | *THADA* | Han Chinese | Keep | 21151128 |
| rs1275468 | 12:75935157 | C | T | 2 | *KRR1* | European | Keep | 26416764 |
| rs13164856 | 5:131813204 | T | C | 3 | *IRF1/RAD50* | European | Keep | 26416764; 30566500 |
| rs13405728 | 2:48978159 | A | G | 2 | *LHCGR* | Han Chinese | Keep | 21151128 |
| rs13429458 | 2:43638838 | A | C | 3 | *THADA* | Han Chinese | Keep | 21151128 |
| rs17186366 | 6:159898261 | C | T | 3 | *SOD2* | Mixed | Keep | 32289280 |
| rs1784692 | 11:113949232 | A | G | 3 | *ZBTB16* | European | Keep | 30566500 |
| rs2059807 | 19:7166109 | G | A | 3 | *INSR* | Han Chinese | Keep | 22885925 |
| rs2178575 | 2:213391766 | A | G | 3 | *ERBB4* | European | Keep | 30566500 |
| rs2268361 | 2:49201612 | C | T | 1 | *FSHR* | Han Chinese | Keep | 22885925 |
| rs2271194 | 12:56477694 | T | A | 2 | *ERBB3/RAB5B* | European | Keep | 30566500 |
| rs2272046 | 12:66224461 | A | C | 2 | *HMGA2* | Han Chinese | Keep | 22885925 |
| rs2349415 | 2:49247832 | T | C | 2 | *FSHR* | Han Chinese | Keep | 22885925 |
| rs2479106 | 9:126525212 | G | A | 1 | *DENND1A* | Han Chinese | Keep | 21151128 |
| rs3802457 | 9:97741336 | G | A | 3 | *C9orf3* | Han Chinese | Keep | 22885925 |
| rs4784165 | 16:52347819 | G | T | 3 | *TOX3* | Han Chinese | Keep | 22885925 |
| rs6022786 | 20:52447303 | A | G | 3 | *SUMO1P1* | Han Chinese | Keep | 22885925 |
| rs705702 | 12:56390636 | G | A | 3 | *RAB5B,SUOX* | Han Chinese | Keep | 22885925 |
| rs804279 | 8:11623889 | T | A | 2 | *GATA4 /NEIL2* | European | Keep | 26284813; 30566500 |
| rs8043701 | 16:52375777 | T | A | 2 | *TOX3* | European | Keep. Using proxy SNP rs11075468 | 30566500 |
| rs9696009 | 9:126619233 | A | G | 1 | *DENND1A* | European | Keep | 30566500 |
| rs10739076 | 9:5440589 | A | C | / | *PLGRKT* | European | Removed because no results in most traits | 30566500 |
| rs113168128 | 2:212291772 | A | G | / | *ERBB4* | Mixed | Removed because no results in most traits | 32289280 |
| rs7563201 | 2:43561780 | A | G | / | *THADA* | European | Removed because no results in most traits | 26416764; 30566500 |
| rs853854 | 20:31420757 | A | T | / | *MAPRE1* | European | Removed because no results in most traits | 30566500 |
| rs7864171 | 9:97723266 | G | A | / | *FANCC* | European | Removed, 1) no results in most traits ;2) in LD with rs10993397 R^2^=0.96, D'=0.98 | 30566500 |
| rs10818854 | 9:126446778 | A | G | / | *DENND1A* | Han Chinese | Removed, in LD with rs10986105, R^2^=0.8, D'=1 | 21151128 |
| rs4385527 | 9:97648587 | G | A | / | *C9orf3* | Han Chinese | Removed, in LD with rs10993397, R^2^=0.96, D'=0.98 | 22885925 |
| rs11031006 | 11:30226528 | A | G | / | *FSHB/KCNA4* | European | Removed, in LD with rs11031006, R^2^=1, D'=1 | 26416764; 26284813 |
| rs1894116 | 11:102070639 | G | A | / | *YAP1* | Han Chinese | Removed, in LD with rs11225154, R^2^=0.93, D'=1 | 22885925 |
| rs1795379 | 12:75941042 | C | T | / | *KRR1* | European | Removed, in LD with rs1275468, R^2^=0.97, D'=1 | 30566500 |
| rs1351592 | 2:213394712 | G | C | / | *ERBB4* | European | Removed, in LD with rs2178575, R^2^=0.9, D'=0.9999 | 26416764 |

Supplementary table 2. List of GWAS datasets of PCOS-related traits and disease outcomes.

| **Trait** | **Dataset ID** | **Sample size** | **Ancestry** | **PMID** |
| --- | --- | --- | --- | --- |
| Fasting Insulin with adj. BMI | ebi-a-GCST005217 | 4,409 | European | 24699409 |
| Fasting Insulin without adj. BMI | ebi-a-GCST005185 | 51,750 | European | 22581228 |
| Fasting glucose | ebi-a-GCST005186 | 58,074 | European | 22581228 |
| HOMA_IR, age, sex adjusted | ebi-a-GCST005179 | 37037 | European | 20081858 |
| BMI-female | ieu-a-974 | 171,977 | European | 25673413 |
| WC-female | ieu-a-62 | 134,593 | Mixed | 25673412 |
| WC-female, BMI adjusted | ieu-a-68 | 134,038 | Mixed | 25673412 |
| WHR-female | ieu-a-74 | 124,591 | Mixed | 25673412 |
| WHR-female, BMI adjusted | ieu-a-80 | 123,302 | Mixed | 25673412 |
| TG | ieu-a-302 | 177,861 | Mixed | 24097068 |
| HDL | ieu-a-299 | 187,167 | Mixed | 24097069 |
| TC | ieu-a-301 | 187,365 | Mixed | 24097070 |
| LDL | ieu-a-300 | 173,082 | Mixed | 24097071 |
| SHBG | ukb-d-30830_raw | 312,215 | European | / |
| LH | prot-a-529 | 3,301 | European | 29875488 |
| FSH | prot-c-3032_11_2 | 996 | European | 28240269 |
| **Outcome** | **dataset id** | **N** | **Ancestry** | **PMID** |
| CAD | ebi-a-GCST005195 | 122733 case, 424528 control | / | 29212778 |
| T2DM | ebi-a-GCST006867 | 61714 case, 593952 control | European | 30054458 |
| Breast cancer | ieu-a-1126 | 122977 case, 105974 control | European | 29059683 |
| ER+ breast cancer | ieu-a-1127 | 69501 case | European | 29059683 |
| ER- breast cancer | ieu-a-1128 | 21468 case | European | 29059683 |
| Ovary cancer | ukb-b-18157 | 1087 case,  461923 control | European | / |
| Endometrial cancer | ukb-b-13545 | 1151 case,  461782 control | European | / |

WC: waist circumference; WHR: waist-hip ratio; ER: estrogen receptor; T2DM: type 2 diabetes mellitus; CAD: coronary artery disease; SHBG: sex hormone binding globulin; BMI: body mass index; LH: luteinizing hormone; HOMA-IR: homeostatic model assessment of insulin resistance; FI: fasting insulin; FG: fasting glucose; HDL: high-density lipoprotein; LDL; low-density lipoprotein; TG: triglyceride; TC: total cholesterol.

Supplementary table 4. The 53 instrumental variables for composed insulin resistant phenotype

| **SNV** | **Gene** | **CHR:BP** | **EA** | **NEA** | **EA frequency** | **β** | **s.e.** |
| --- | --- | --- | --- | --- | --- | --- | --- |
| rs17386142 | *DMRTA2* | 1:50815783 | C | T | 0.92744 | 0.02243026 | 0.00400815 |
| rs11577194 | *CSF1* | 1:110500175 | T | C | 0.4789 | 0.01351333 | 0.00196299 |
| rs9425291 | *DNM3* | 1:172312769 | A | G | 0.4288 | 0.01493736 | 0.00199413 |
| rs4846565 | *RNU5F-1 / LYPLAL1* | 1:219722104 | G | A | 0.6939 | 0.01628592 | 0.00208385 |
| rs2249105 | *CEP68* | 2:65287896 | A | G | 0.6227 | 0.0159435 | 0.00201374 |
| rs492400 | *USP37* | 2:219349752 | T | C | 0.6029 | 0.01295845 | 0.00203774 |
| rs308971 | *SYN2 / PPARG* | 3:12116620 | G | A | 0.1385 | 0.0236288 | 0.00291537 |
| rs3864041 | *COL6A4P1* | 3:15185634 | T | C | 0.6227 | 0.01093161 | 0.00212383 |
| rs9881942 | *ADCY5* | 3:123082416 | A | G | 0.4354 | 0.01271202 | 0.00197336 |
| rs6822892 | *PDGFC* | 4:157734675 | A | G | 0.6464 | 0.01815056 | 0.00208385 |
| rs4976033 | *PIK3R1* | 5:67714246 | G | A | 0.3799 | 0.01686784 | 0.00215357 |
| rs6887914 | *MCC* | 5:112711486 | C | T | 0.781 | 0.01435025 | 0.00246364 |
| rs1045241 | *TNFAIP8* | 5:118729286 | C | T | 0.7427 | 0.01367552 | 0.00223379 |
| rs2434612 | *EBF1* | 5:158022041 | G | A | 0.2071 | 0.01682953 | 0.00248483 |
| rs966544 | *CPEB4* | 5:173350405 | G | A | 0.3074 | 0.01394734 | 0.00214223 |
| rs12525532 | *ANKS1A* | 6:35004819 | T | C | 0.3958 | 0.01488394 | 0.00203393 |
| rs9492443 | *L3MBTL3* | 6:130398731 | C | T | 0.7639 | 0.01439165 | 0.00223379 |
| rs17169104 | *MEOX2* | 7:15883727 | G | C | 0.3417 | 0.0183302 | 0.00250369 |
| rs4738141 | *EYA1* | 8:72469742 | G | A | 0.2493 | 0.01737655 | 0.00249145 |
| rs498313 | *MIR548H3* | 9:78034169 | A | G | 0.6913 | 0.01229841 | 0.00212383 |
| rs11231693 | *MACROD1* | 11:63862612 | A | G | 0.06069 | 0.0315533 | 0.00423559 |
| rs17402950 | *ATF7IP* | 12:14571671 | G | A | 0.05541 | 0.03115788 | 0.00580273 |
| rs718314 | *ITPR2* | 12:26453283 | G | A | 0.2348 | 0.0160844 | 0.00226414 |
| rs7323406 | *ANKRD10* | 13:111628195 | A | G | 0.2784 | 0.0150323 | 0.00303812 |
| rs7176058 | *C15orf54* | 15:39464167 | A | G | 0.8364 | 0.01487385 | 0.00263426 |
| rs8032586 | *LOC100287559* | 15:73081067 | C | T | 0.8813 | 0.02129746 | 0.0042999 |
| rs754814 | *ZMYND15* | 17:4657034 | T | C | 0.7309 | 0.01174158 | 0.00219418 |
| rs6066149 | *EYA2* | 20:45602638 | G | A | 0.7612 | 0.0138085 | 0.00226414 |
| rs683135 | *MACF1* | 1:39895460 | A | G | 0.2678 | 0.01908287 | 0.00216275 |
| rs10195252 | *COBLL1 / GRB14* | 2:165513091 | T | C | 0.5818 | 0.02718435 | 0.00205301 |
| rs2943645 | *IRS1* | 2:227099180 | T | C | 0.6227 | 0.03059101 | 0.00201374 |
| rs295449 | *KLHL18* | 3:47375955 | A | G | 0.595 | 0.01443483 | 0.00230587 |
| rs11130329 | *TMEM110-MUSTN1* | 3:52896855 | A | C | 0.8641 | 0.02284733 | 0.00400104 |
| rs645040 | *MSL2* | 3:135926622 | T | G | 0.7691 | 0.02504529 | 0.00239352 |
| rs2699429 | *DOK7* | 4:3480136 | C | T | 0.4063 | 0.01647976 | 0.00208385 |
| rs3822072 | *FAM13A* | 4:89741269 | A | G | 0.4881 | 0.02125998 | 0.00199413 |
| rs4865796 | *ARL15 / FST* | 5:53272664 | A | G | 0.7071 | 0.01584572 | 0.00212383 |
| rs459193 | *ANKRD55* | 5:55806751 | G | A | 0.715 | 0.02218311 | 0.00223379 |
| rs6937438 | *LOC100132354* | 6:43815364 | A | G | 0.7084 | 0.01497735 | 0.00219418 |
| rs2745353 | *RSPO3* | 6:127452935 | T | C | 0.4789 | 0.0183932 | 0.00197336 |
| rs3861397 | *LOC645434* | 6:139828916 | G | A | 0.3417 | 0.02056984 | 0.00205301 |
| rs972283 | *KLF14* | 7:130466854 | G | A | 0.5435 | 0.02284548 | 0.00212971 |
| rs2126259 | *PPP1R3B* | 8:9185146 | T | C | 0.09103 | 0.04425118 | 0.00315658 |
| rs1011685 | *LPL* | 8:19830769 | C | T | 0.8773 | 0.11616272 | 0.00326565 |
| rs7005992 | *TRIB1* | 8:126528955 | C | G | 0.1359 | 0.01791116 | 0.00286476 |
| rs10995441 | *NRBF2* | 10:64869239 | G | T | 0.2361 | 0.01627273 | 0.00239498 |
| rs7973683 | *CCDC92 / DNAH10* | 12:124449223 | C | A | 0.6385 | 0.02415614 | 0.00205301 |
| rs7227237 | *LIPG* | 18:47174679 | C | T | 0.7678 | 0.01780009 | 0.00321892 |
| rs8101064 | *INSR* | 19:7293119 | T | C | 0.04354 | 0.0565322 | 0.00816483 |
| rs4804833 | *MAP2K7* | 19:7970635 | A | G | 0.4103 | 0.01772949 | 0.00212833 |
| rs4804311 | *MYO1F* | 19:8615589 | A | G | 0.8905 | 0.03672984 | 0.00358621 |
| rs731839 | *PEPD* | 19:33899065 | G | A | 0.3417 | 0.02333592 | 0.0020912 |
| rs132985 | *PLA2G6* | 22:38563471 | C | T | 0.5646 | 0.01759948 | 0.00194317 |

SNV: single nucleotide variant; EA: effect allele. NEA: non-effect allele; s.e.: standard error.
